# A Single Drop of Fingerstick Blood for Quantitative Antibody Response Evaluation After SARS-CoV-2 Vaccination

**DOI:** 10.1101/2021.04.11.21255278

**Authors:** Jinwei Du, Dayu Zhang, Hannah May, Yulia Loginova, Eric Chu, Roberta Madej, Chuanyi M. Lu, Joseph A. Pathakamuri, Daniel Kuebler, Jocelyn V. Neves, Aiguo Zhang, Michael Y. Sha

## Abstract

Among several COVID vaccines that have been approved, the Moderna and Pfizer-BioNTech vaccines are mRNA vaccines that are safe and highly effective at preventing COVID-19 illness. Studies have demonstrated that neutralizing antibody responses elicited by these vaccines correlate strongly with antibodies measured by immunoassays such as ELISA. To monitor the antibody level duration of vaccine-induced immune responses in vaccinated population, cost-effective and easily implementable antibody testing methodologies are urgently needed. In this study, we evaluated the feasibility of using a single drop of fingerstick blood collected with flocked swabs for a high-throughput and quantitative anti-SARS-CoV-2 spike (S1) IgG antibody immunoassay. A total of 50 voluntary subjects participated and donated fingerstick blood samples before and after receiving the Moderna mRNA vaccine. Among all individuals tested, no anti-SARS-CoV-2 S1 IgG antibody was detected before vaccination and on day 7 after receiving the first vaccine dose. On day 14 after the first dose, a significant amount of anti-SARS-CoV-2 S1 IgG antibody was detected in all participants’ samples. By the end the third week from the first dose, the median anti-SARS-CoV-2 S1 IgG concentration increased to 44.9 ug/mL. No anti-SARS-CoV-2 nucleocapsid (N) protein IgG antibody was detected in any of the participants during the study period, indicating that the anti-SARS-CoV-2 S1 IgG assay is specific for the mRNA vaccine induced antibodies.Comaprison of venous blood plasma and fingerstick blood for anti-SARS-CoV-2 S1 IgG shown a higher correlation. Furthermore, the fingerstick blood dried swab samples are stable for at least 4 days. In summary, we demonstrated that a single drop of fingerstick blood collected with flocked swab can be used for quantitative detection and monitoring of anti-SARS-CoV-2 spike IgG responses after receiving COVID-19 vaccination. This testing platform does not require venous blood draw and can be easily implemented for large scale antibody testing in vaccinated populations.

## Introduction

COVID-19, caused by infection with the novel severe acute respiratory syndrome coronavirus-2 (SARS-CoV-2), is associated with a spectrum of clinical manifestations ranging from asymptomatic infection, minor flu-like illness to acute respiratory distress syndrome (ARDS), severe pneumonia and death. SARS-CoV-2 is a single-stranded RNA virus with high sequence identity to the SARS-CoV. The genome sequence of SARS-CoV-2 displays 50% homologous to MERS-CoV, 79% with SARS-CoV, 88% with two bat-derived SARS-like coronaviruses ((Bat-SL-CoVZC45 and Bat-SL-CoVZXC21),) and 96% with Bat-SARSr-CoV RaTG13 ^1^.

Antibodies that bind to the SARS-CoV-2 spike protein have been shown the potential in blocking the viral entry into cells in vitro and appear to play a key role in the protective immune responses to SARS-CoV-2 infection ^2-6^. Safe and effective COVID-19 vaccines have been developed and distributed, and millions of people in the United States have received vaccination. Among the FDA approved COVID-19 vaccines, the Pfizer BioNTech (BNT162b1 and BNT162b2) and Moderna vaccines (mRNA-1273) are lipid nanoparticle–encapsulated mRNA vaccines and the Janssen vaccine is an adenovirus (AD26) vector-based vaccine ^7^. Krammer et al ^8^ showed majority of seronegative participants had variable and relatively low SARS-CoV-2 IgG responses within 9 to 12 days after first does of vaccination (439 AUC at 9 to 12 days; 1016 AUC at 13 to 16 days; 1037 AUC at 17 to 20 days; 1293 AUC at 21 to 27 days; and 3316 AUC after the second dose). Further their studies showed individuals with preexisitng immunity receiving a single dose of mRNA vaccine elicitng a rapid immune responses 10 to 45 times as high antibodies titers compared to those without pre-exisitng immunity. The median antibody titers in preexisitng immunity group were similar to or exceeded titers found in seronegative participants who received two vaccinations ^8^. In similar studies conducted on Astra Zeneca AZD1222 a adenovirus-vectored vaccine (formerly ChAdOx1 nCoV-19), Folegatti et al reported that Anti-spike IgG responses rose by day 28 (median 157 ELISA units or [EU]) and were boosted following a second dose (639 EU). In addition, neutralizing antibody responses against SARS-CoV-2 were detected in 100% participants when measured in PRNT50, indicating that neutralising antibody responses in vaccinated individuals correlated strongly with antibody responses measured by ELISA^9^.

In a related study results suggest that a standardized ELISA based antibody test might be sufficient to predict protection following vaccination. Ramasamy et al ^10^ found that in participants who received two doses of vaccine, the median anti-spike SARS-CoV-2 IgG responses measured by ELISA were elevated 28 days after the boost dose across the three age-based cohorts (18–55 years, 20713 AU/mL; 56–69 years, 16170 AU/mL, and ≥70 years 17561 AU/mL). Richmond et al ^11^ also confirmed strong correlations between binding/blocking IgG antibodies by ELISA and neutralizing antibodies by microneutralization assay after receiving a spike (S)-protein (S-Trimer) based vaccine. Indeed, consistent with the high IgG and neutralizing antibody titers, a single dose of mRNA-RBD conferred near-complete protection against SARS-CoV-2 infection *in hACE2 transgenic mice* ^12^. In patients infected with SARS-CoV-2, the IgG antibodies to SARS-CoV-2 RBD were strongly correlated with anti-S neutralizing antibody titers. These titers demonstrated little to no decrease over 75 days symptom onset ^13^.

Recent real-word condition studies on health care personnel, first responders, and other essential and frontline workers confirmed mRNA COVID-19 vaccines (Pfizer-BioNTech’s BNT162b2 and Moderna’s mRNA-1273) are highly effective, 90% with full immunization and 80% with partial full immunization ^14^. Studies on BNT162b2 mRNA vaccine effectiveness in UK healthcare workers demonstrated significant protection from asymptomatic and symptomatic infection with protection increasing from day 10 onwards, and plateauing after 21 days after first dose ^15^. Therefore, the anti-SARS-CoV-2 spike (S) specific IgG antibodies measured by ELISA could be used to predict protection after SARS-CoV-2 infection or COVID-19 vaccination based immunity.

Notably, Karp et al ^16^ measured finger-prick dried blood spot based SARS-CoV-2 antibody assay and demonstrated that results of dried fingerstick blood correlated well with that of paired plasma samples. Iyer et al ^17^ further confirmed that anti-RBD IgG dried blood spots (DBS) measurements had a high degree of linear correlation with plasma. As more vaccines are being made available for large scale vaccination, it is highly desirable to have a simple and convenient test for detecting anti-SARS-CoV-2 IgG antibody. In this study, we evaluated the feasibility of using a single drop of fingerstick blood sample collected with nylon flocked swabs for an quantitative detection of anti-SARS-CoV-2 S1 IgG antibody, and demonstrated that fingerstick blood is sufficient for quantitative measurement of the antibody responses after receiving a mRNA COVID-19 vaccine.

## Materials and Methods

### Fingerstick blood study design and ethics

Deidentified fingertip blood samples were used in the study. All fingerstick blood specimens were collected in January – March 2021 and were tested at DiaCarta’s CLIA laboratory. Other than anti-SARS-CoV-2 S1 IgG results (positive or negative), no other information were included in study analysis and no patient clinical chart reviews were performed. This study was approved by the Institutional Review Board (IRB) at Franciscan University of Steubenville (IRB #2021-05) as a no-subject contact study with waiver of consent and as exempt under category 4.

### Fingerstick blood dried swab collection

Fingerstick blood dried swab collection kit contains disposable lancets, alcohol pads, a swab, one 1 mL microtube and instruction sheet. First, clean the desired puncture site using alcohol prep pad. Remove the cap of safety lancet and stick the fingerstick with lancet. Squeeze fingerstick to obtain one drop of blood (approx. 20µL). Next, wipe the blood onto the stick of nylon flocked swab (Kangjian, Jiangsu, China). Lastly, place the swab into screw cap tube, break swab at the score mark, and leave the stick of swab in screw cap tube. A guidance diagram show in Supplementary Figure 1.

### SARS-CoV-2 S1 & N protein and Reagents

The recombinant SARS-CoV-2 Spike protein S1 (RBD, His Tag) containing 330-524 amino acids of Spike protein and the SARS-CoV-2 nucleocapsid (N) protein (PMC 827, NP2) were produced from HEK293 suspension cells (ProMab Biotechnologies Inc, CA). SARS-CoV-2 Spike S1 Antibody (human chimeric, IgG isotype) was purchased from GenScript Biotech Corporation (Piscataway, NJ). PE conjugated anti-human IgG Fc antibody was purchased from BioLegend (San Diego, CA). MagPlex Microsphere and xMAP^®^ Antibody Coupling (AbC) kit were purchased from Luminex (Austin, TX). Hemoglobin (human), bilirubin and EDTA were purchased from Sigma-Aldrich (St. Louis, MO).

### Detection of anti-SARS-CoV-2 Spike IgG from fingerstick blood dried swab

First, 300uL of PBS-1% BSA buffer was added to the 1mL tube that contained one drop of fingerstick blood in dried swab and waited for 5 minutes. Then briefly vortex the tube to release the blood into the buffer. The inoculated buffer is used for IgG testing. Principle of the assay was shown in Figure 1. Recombinant spike protein 1 (S1) RBD was covalently coupled to the surface of MagPlex^®^ Microspheres (magnetic beads) via a carbodiimide linkage using xMAP^®^ Antibody Coupling (AbC) kit. S1 RBD protein coated magnetic beads and human specimens were mixed and incubated at room temperature for 1.0 hour. If present, the IgG antibodies in specimen against S1 RBD protein (the antigen) will bind to the coated magnetic beads. After washing, PE conjugated anti-human IgG antibody was added to the reaction mixture and incubated at room temperature for 0.5 hours. After another washing, PE fluorescence of each well in a 96-well microplate was measured on MAGPIX^®^ instrument for Median Fluorescence Intensity (MFI). Interpretation of the testing results was performed by calculating the MFI ratio of each sample to the average MFI of two blank wells included in each test run.

**Figure 1.**
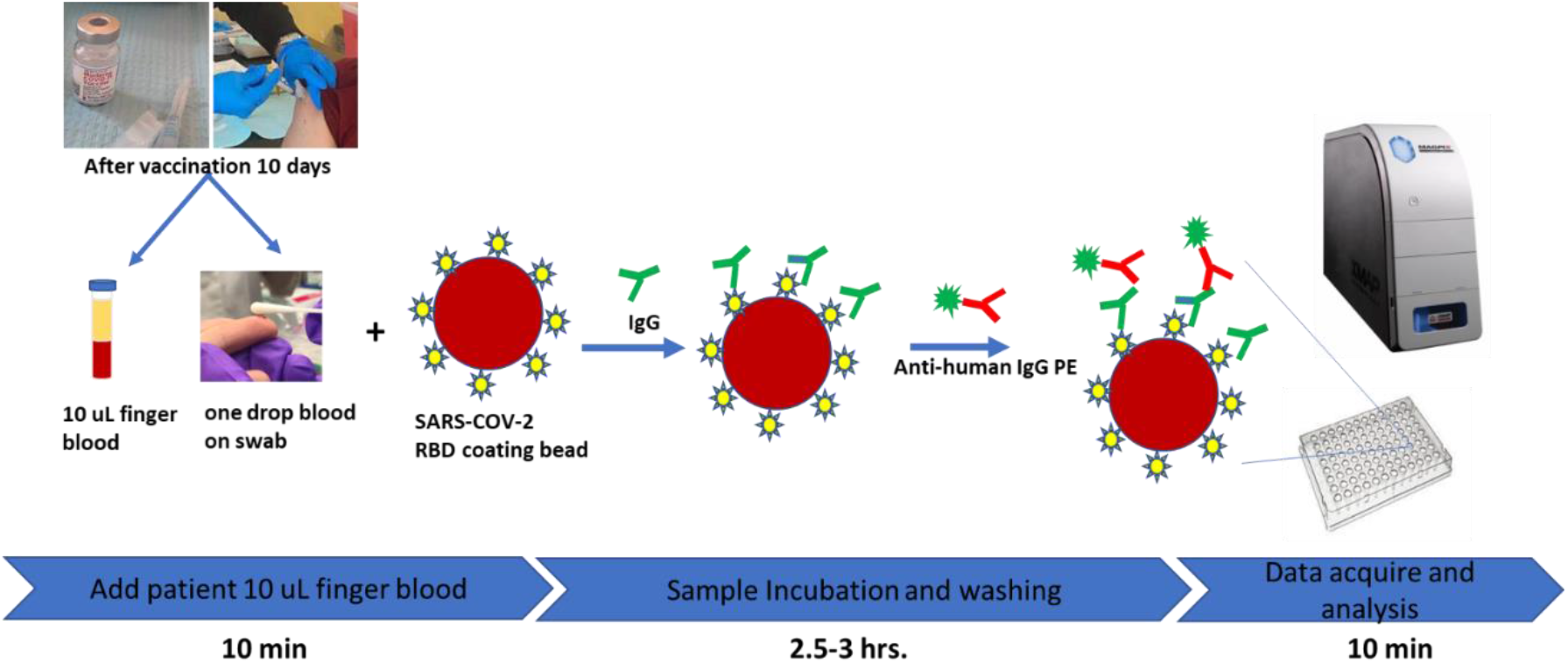
A high throughput immunoassay platform for anti-SARS-CoV-2 S1 IgG detection using a single drop of fingerstick blood collected in EDTA tube or on flocked swab.

## Results

### Development of a quantitative immunoassay for anti-SARS-CoV-2 S1 protein IgG antibody

Previously, we reported the development of a semi-quantitative immunoassay for anti-SARS-CoV-2 IgG ^18^ which showed that Positive percent agreement (PPA) was 46.15%, 61.54%, and 97.53% for samples collected on 0-7 days, 8-14 days, and ≥15 days from symptom onset, respectively, and Negative Percent Agreement (NPA) was 98.23%. No cross-reactivity was observed to patient samples positive for IgG antibodies against the following pathogens: *HIV, HAV, HBV, RSV, CMV, EBV, Rubella, Influenza A*, and *Influenza B*. In order to evaluate the post-vaccination immune responses, we aimed to develop a quantitative immunoassay. SARS-CoV-2 Spike S1 Antibody (HC2001) obtained from Genscript Biotech Corporation (Catalog number A02038) was serially diluted with PBS-1%BSA buffer to prepare five standards with various concentrations: 40, 10, 2.5, 0.62, 0.16 µg/mL. A quantitative measurement was performed using Logistic 5P Weighted analysis method on Luminex xPONENT (version 4.2.1705.0) software. A typical standard curve was shown in Supplementary Figure 2. Using this calibration curve, we can calculate the IgG concentration in a specimen through its Median Fluorescence Intensity (MFI) value.

### Anti-SARS-CoV-2 S1 IgG Antibody Responses After SARS-CoV-2 mRNA Vaccine

Since venous blood could be logistically challenging to collect and process during the ongoing pandemic, we evaluated the feasibility of in-home collection of fingerstick blood, followed by shipping to a central laboratory for anti-SARS-CoV-2 IgG antibody detection. First, one drop of fingerstick blood (approximately 20µL) was collected with a swab following the instructions depicted in Supplementary Figure 1. Before testing, the dried swab was immersed in 300µL PBS-1% BSA buffer for 5 minutes, followed by brief vortex of the tube was quickly vortexed to release blood into the buffer. 50µL inoculated buffer was used for IgG antibody testing by incubating it with Luminex microspheres coated with SARS-CoV-2 S1 RBD protein at room temperature for 1 hour followed by incubation with anti-human IgG antibody (PE conjugated). Data acquisition was then performed on the samples using a Luminex MAGPIX reader in a 96-well format (Figure 1).

Using the aforementioned method, we tested anti-SARS-CoV-2 S1 IgG in fingerstick blood collected from donors without history of COVID-19 as well as those with a previous COVID-19 infection as confirmed by an FDA-EUA approved RT-PCR kit (QuantiVirus™ SARS-CoV-2 Test Kit, DiaCarta Inc, CA). As shown in Table 1, 30 subjects were tested before receiving the first dose of the Moderna COVID-19 mRNA vaccine, and all tested negative for anti-SARS-CoV-2 spike IgG. A total of 56 samples were collected and tested at different time points after vaccination. Two weeks after the first-dose of vaccine, 25 of 28 subjects tested positive for SARS-CoV-2 S1 IgG in this assay. S1 IgG detection rate was 89.3%. Interestingly, all 3 negative are elderly individuals over 80 years of age. By three weeks after the post first dose of the vaccination, all the subjects tested positive for anti-SARS-CoV-2 S1 IgG, detection rate was 100%. Among the subjects tested, we further quantitatively analyzed the SARS-CoV-2 S1 IgG testing results of 15 individuals (age: 22∼58 years, median 35 years; 10 males, 5 females). As shown in Table 2, no anti-SARS-CoV-2 S1 IgG antibody was detected before vaccination and on day 7 post first dose of vaccine (Moderna). By the ends of the second week and the third week after receiving the first dose, a significant amount of SARS-CoV-2 IgG antibody was detected in all individuals. The lowest levellevel detected was 0.54 µg/mL, while the highestlevels detected were 59.01 µg/mL, with the median being 20.78 µg/mL of IgG antibody. This data indicates that the fingerstick blood-based testing method is sufficient for detecting and monitoring the development of anti-SARS-CoV-2 Spike IgG antibody after vaccination.

**Table 1.** Fingerstick blood-based testing of anti-SARS-CoV-2S1 IgG after COVID-19 vaccination

| **Group** | **Number of subjects** | **IgG Positive (Detection Rate%)** |
| --- | --- | --- |
| Pre-vaccination | 30 | 0 (0.0%) |
| 2 weeks post 1st dose vaccination | 28 | 25 (89.3%) |
| 3 weeks post 1st dose vaccination | 17 | 17 (100%) |
| 1 week post 2nd dose vaccination | 11 | 11 (100%) |

**Table 2.** Fingerstick blood-based testing of anti-SARS-CoV-2 S1 IgG after COVID-19 vaccination

| Sample ID | Anti-SARS-CoV-2 S1 IgG (µg/mL) |  |  |  |
| --- | --- | --- | --- | --- |
|  | Pre-Vaccination | 7-day | 14-day | 21-day |
| DIA_01 | Undetectable | Undetectable | 2.2 | 48.16 |
| DIA_02 | Undetectable | Undetectable | 2.53 | n/a |
| DIA_03 | Undetectable | Undetectable | 3.00 | 56.89 |
| DIA_04 | Undetectable | Undetectable | 9.93 | 42.59 |
| DIA_05 | Undetectable | Undetectable | 2.80 | n/a |
| DIA_06 | Undetectable | Undetectable | 2.07 | n/a |
| DIA_07 | Undetectable | Undetectable | 2.37 | 45.72 |
| DIA_08 | Undetectable | Undetectable | 44.2 | 59.01 |
| DIA_09 | Undetectable | Undetectable | 8.91 | 44.97 |
| DIA_10 | Undetectable | Undetectable | 2.18 | 44.88 |
| DIA_11 | Undetectable | Undetectable | 0.85 | n/a |
| DIA_12 | Undetectable | Undetectable | 1.93 | 2.74 |
| DIA_13 | Undetectable | n/a | 0.89 | 10.29 |
| DIA_14 | Undetectable | n/a | 36.7 | 43.06 |
| DIA_15 | Undetectable | n/a | 0.54 | n/a |
| Mean |  |  | 20.78 |  |

n/a: data not available

It was noted that, although the median value of IgG concentration in individuals 2 weeks after the first dose of vaccine was similar to those in asymptomatic individuals without known exposure to SARS-CoV-2 but confirmed by RT-qPCR late, on week 3 post vaccination, the median value of IgG level was remarkably increased (2.2 µg/mL, 2.8 µg/mL and 44.9 µg/mL for 1, 2 and 3 weeks, respectively. see Figure 2), indicating that COVID-19 mRNA vaccine is more efficient in inducing IgG antibody responses than asymptomatic SARS-CoV-2 infection.

**Figure 2.**
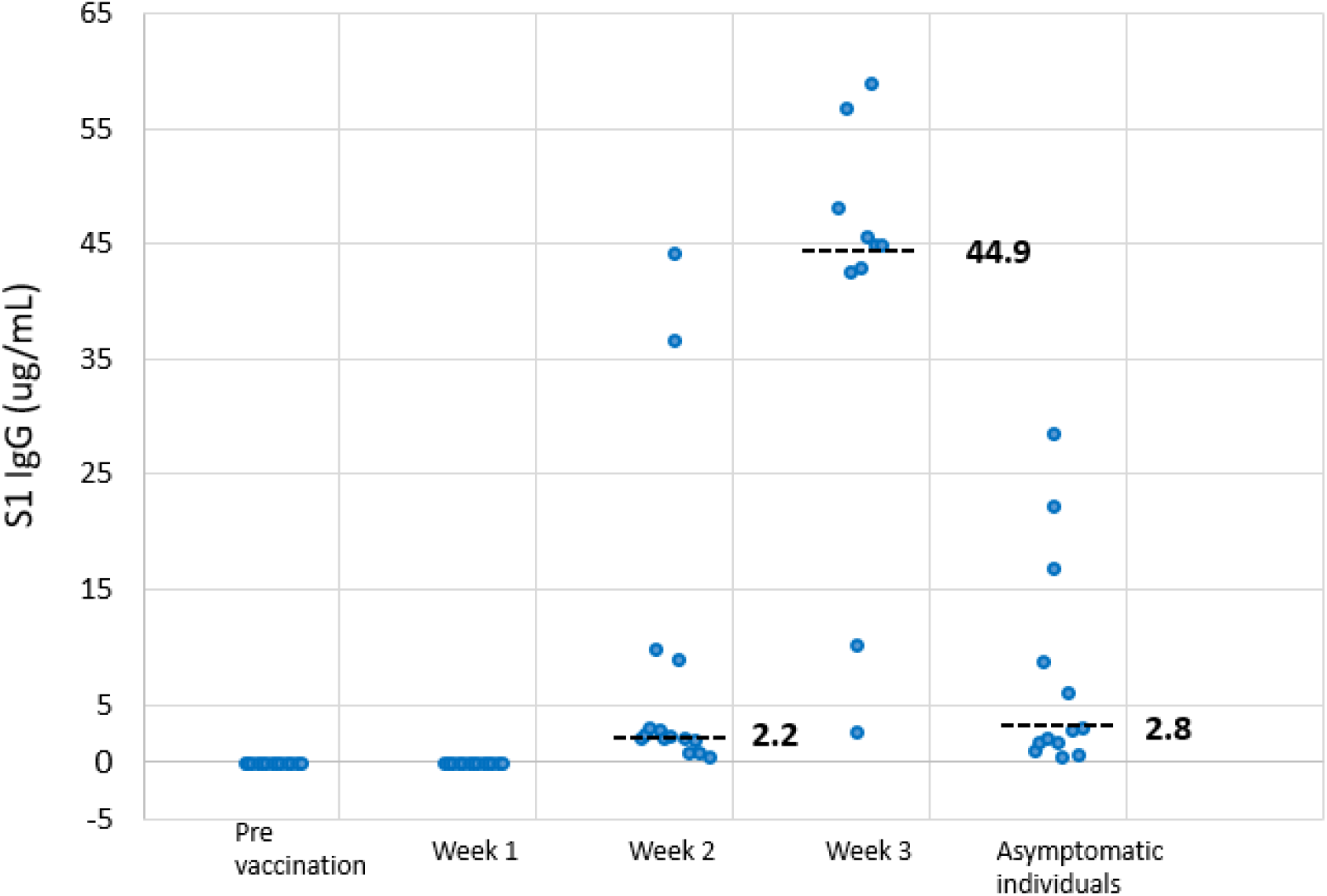
Quantitative measurements of anti-SARS-CoV-2 spike IgG before and after receiving the Moderna COVID-19 mRNA vaccine. The dashed lines and numbers are the median values for each sample groups

Notable, second dose boost the anti-SARS-CoV-2 S1 IgG from average 8525 MFI (3 weeks) up to 10752 MFI in the first week (Figure 3). Gradually, the average IgG increases to 13383 MFI in 4 weeks. It indicates that anti-SARS-CoV-2 S1 IgG achieves a high levele after two months vaccination.

**Figure 3.**
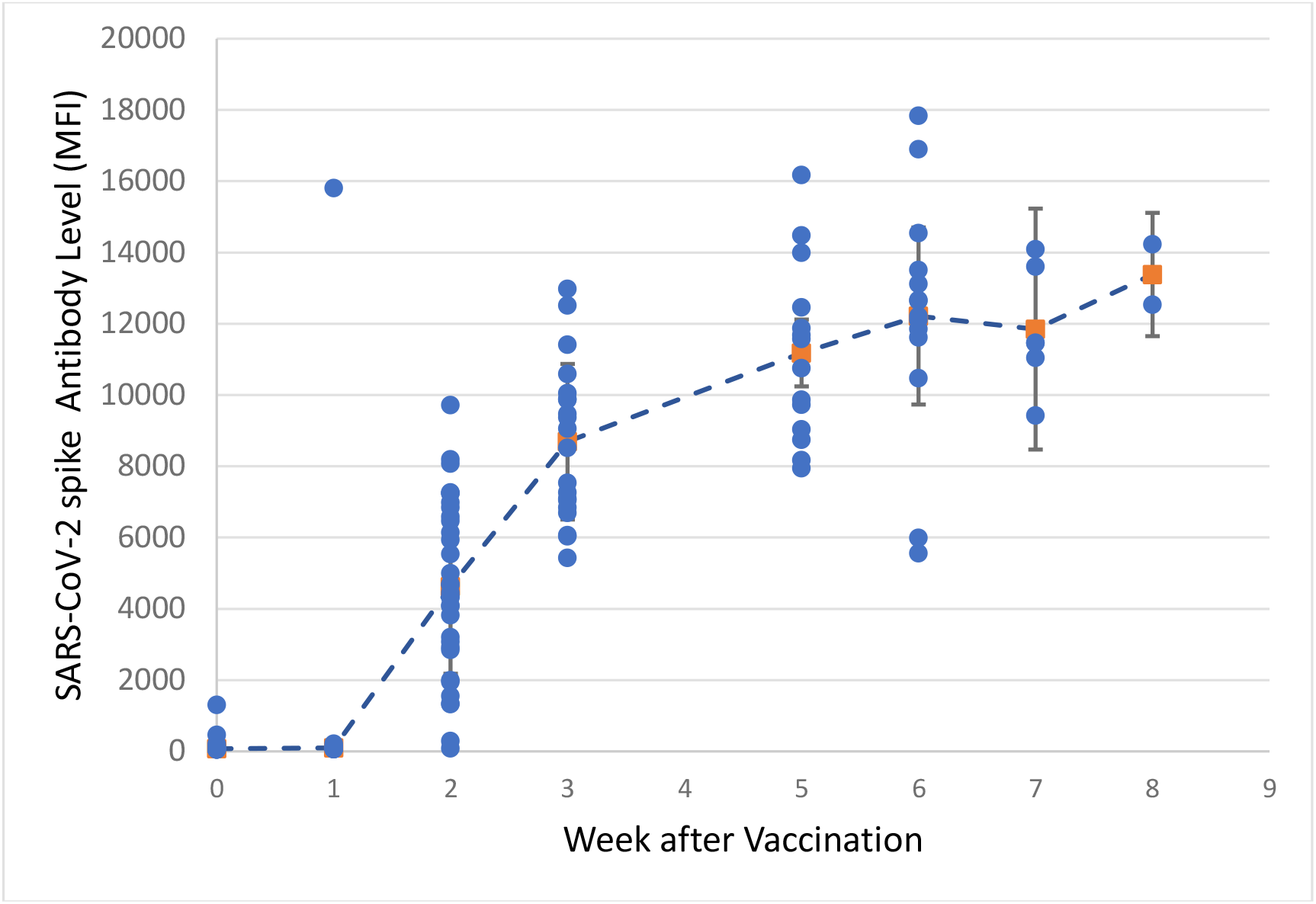
SARS-CoV-2 mRNA vaccine induced humoral immune responses. Shown are the quantitative anti-SARS-CoV-2 S1 IgG antibody levels from 50 vaccinated participants.

### Comparison of fingerstick blood samples with EDTA tube versus on dried swab samples

We examined the relationship between IgG levels detected in fingertip blood samples either stored in EDTA tube or onto flocked swab. Five of paired fingerstick blood samples were tested in parallel. The MFI signals of each paired samples from vaccinated donators were compared side by side. Overall, the samples with EDTA showed higher signal than the dried swab samples. Nonetheless, between the two sample types, the signals are highly correlated with an R^2^ of 0.98 (Figure 4).

**Figure 4.**
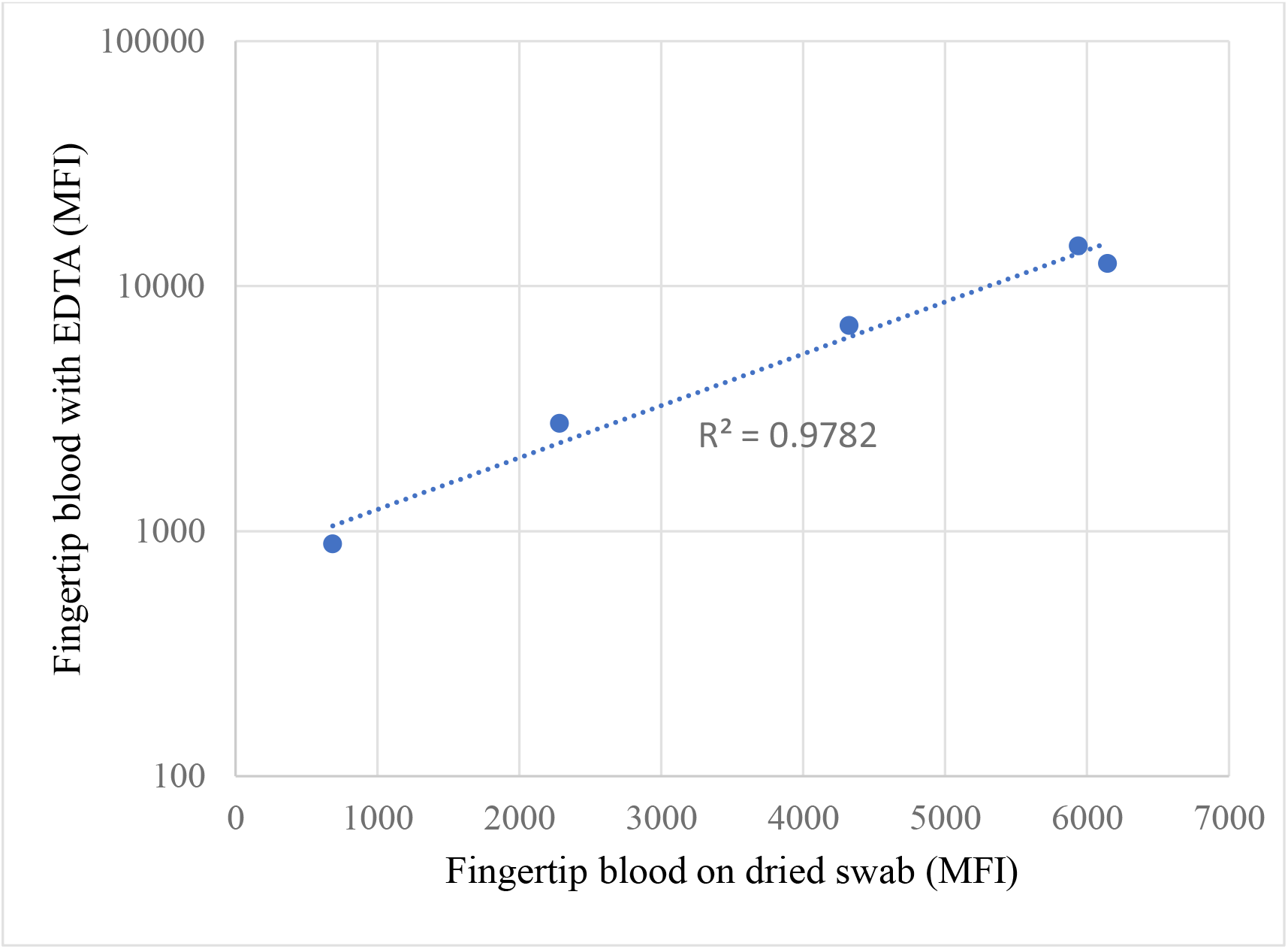
Comparison of fingerstick blood samples collected with EDTA tube and on dry swab

### Highly correlation of venous plasma samples and fingerstick blood dried swab samples

We also compared the ratio between the signal intensity from pairs of venous plasma samples and dried swab samples which it came from vaccinated donators. Shown in Table 3 is the side by side comparison between the two sample type of five pair of vaccinated donators. In average, the MFI in plasma samples are about 4 time higher than that of fingerstick blood dried swab samples. The higher signals in plasma samples are expected as plasma obtained from venous blood contains a higher concentration of IgG antibodies than the more diluted fingerstick whole blood. Unsurprisingly, a highly correlation between plasma and fingerstick blood dried swab were observed (Figure 5).

**Table 3.** Comparision between venous plasma sample and fingerstick blood dried swab sample

| **Sample ID** | **Plasma (MFI)** | **Fingerstick blood (MFI)** | **Ratio** |
| --- | --- | --- | --- |
| DIA001K | 23884 | 6628 | 3.6 |
| DIA002L | 25636 | 6186 | 4.1 |
| DIA003M | 8937 | 2068 | 4.3 |
| DIA004N | 22048 | 4308 | 5.1 |
| DIA005O | 25082 | 8239 | 3.0 |

\* Each pair of venous blood plasma and fingerstick blood came from same donator.

**Figure 5.**
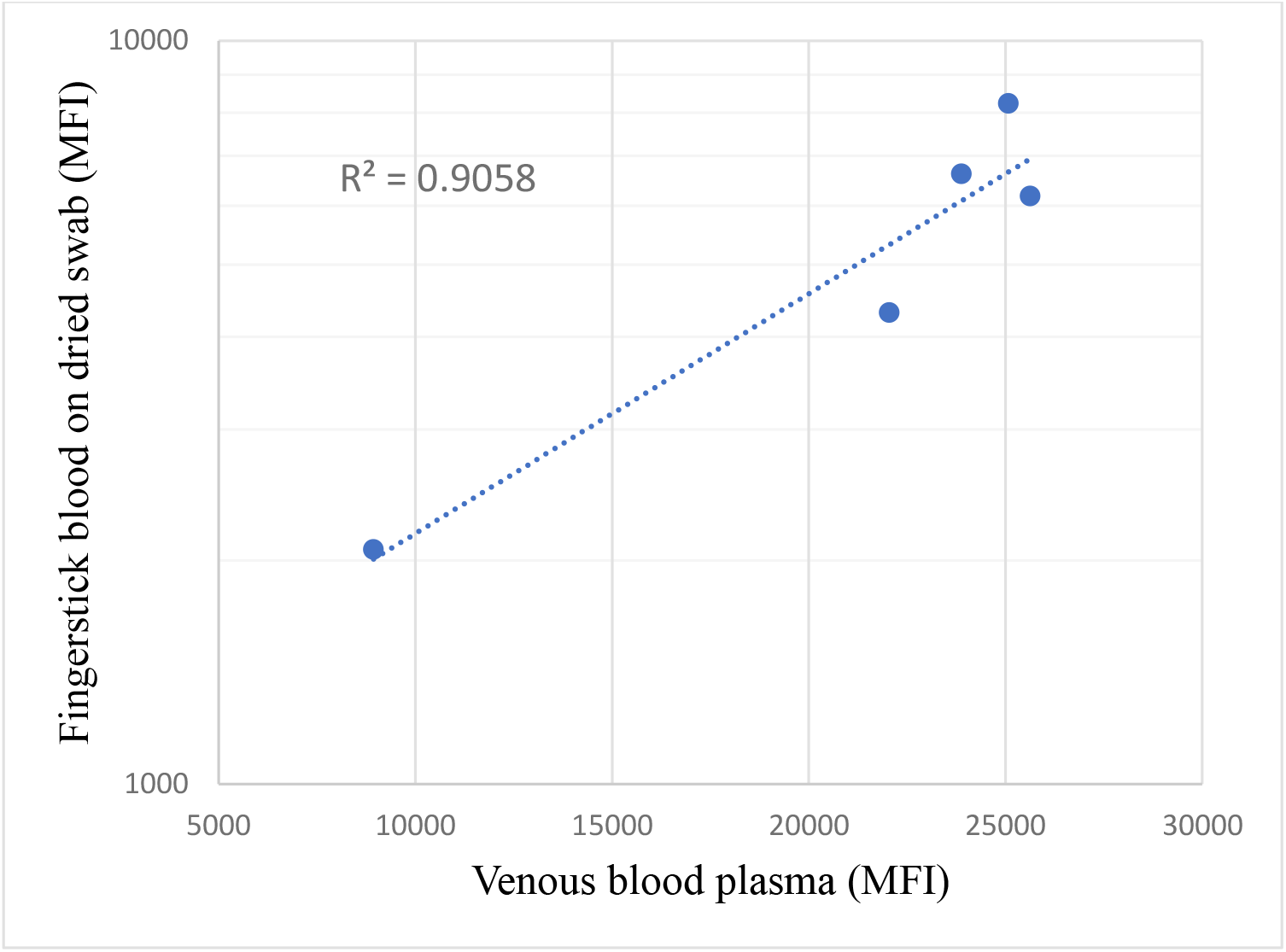
Comparison between venous blood plasma in EDTA tube and fingerstick blood on dried swab.

### mRNA Vaccine elicited anti-SARS-CoV-2 S1 IgG distinguished with infectetion caused anti-SARS-CoV-2 N protein IgG

To affirm the reported MFI signals were indeed specific for the SARS-CoV-2 Spike protein IgG antibodies elicited by mRNA vaccine, five samples were also tested using microspheres conjugated with SARS-CoV-2 nucleocapsid protein (N protein) and SARS-CoV-2 Spike RBD protein coupled microspheres separatly. As shown in Table 4, for these five samples from individuals vaccinated with Moderna SARS-CoV-2 mRNA vaccine, Spike RBD protein coupled microspheres detected high signal with the ratio greater than 200 (the cutoff value is 13.2) while the signal generated by N protein coupled microspheres was close to the background.

**Table 4.** Fingerstick blood sample-based antibody immunoassay using SARS-CoV-2 spike RBD protein versus nucleocapsid protein coupled microspheres

| Sample ID | Ratio of MFI Sample to MFI Background |  |
| --- | --- | --- |
|  | Spike RBD protein coupled microspheres | N protein coupled microspheres |
| DIAA | 252.0 | 1.4 |
| DIAB | 248.2 | 2.7 |
| DIAC | 273.4 | 3.2 |
| DIAD | 227.2 | 1.7 |
| DIAE | 232.7 | 2.1 |

### Stability of fingerstick blood in dried swab

To test the stability of fingerstick blood collected using nylon flocked swabs and stored at room temperature (20±5 °C), total 7 of dried swab samples were tested on day 1, day 2, and day 4 at room temperature. As shown in Table 5, the samples were stable for at least 4 days at room temperature.

**Table 5.** Stability evaluation of fingerstick blood dries swab for anti-SARS-CoV-2 IgG immunoassay

| Sample ID | Day 1 |  | Day 2 |  | Day 4 |  |
| --- | --- | --- | --- | --- | --- | --- |
|  | MFI* | Interpretation | MFI | Interpretation | MFI | Interpretation |
| Dia161 | 4342 | positive | 4241 | positive | n/a | n/a |
| Dia152 | 7771 | positive | 7533 | positive | n/a | n/a |
| Dia151 | 11345 | Positive | n/a | n/a | 11680 | Positive |
| Dia150 | 4689 | Positive | n/a | n/a | 2425 | Positive |
| Dia149 | 10752 | Positive | n/a | n/a | 11004 | Positive |
| Dia148 | 433 | Negative | n/a | n/a | 351 | Negative |
| Dia147 | 68 | Negative | n/a | n/a | 80 | Negative |

\*MFI stands for Median Fluorescence Intensity

## Discussion

We have demonstrated that an at home fingerstick blood collected on flocked dry swab is comparable to fingerstick blood collected into EDTA tube for anti-SARS-CoV-2 Spike IgG detection after full vaccination. A single drop of fingerstick blood with dried swab also has higherly correlation with venous plasma specimen for evoluation of anti-SARS-CoV-2 S1 IgG level. This approach is more convenient for end users, because there is no need for users to visit healthcare facility or a diagnostic laboratory. Furthermore, a singl drop of fingerstick blood was successfully applied to a quantitative anti-SARS-CoV-2 spike IgG assay, which can be used for monitoring the antibody response status overtime after full vaccination with COVID mRNA vaccine.

Our study indicated that 3 weeks after receiving the first dose of mRNA vaccine, a significant amount of anti-SARS-CoV-2 S1 IgG antibody can be detected in most individuals at an average of 44.9 ug/mL, comparedto their basilines before vaccination; with the execption of three people who are over 80 years old. This result is consistent in concordance with a recent report that after of vaccination the antibody response was increased up to 1016 AUC on days 13-16, compared to 1 AUC on days 0-4 days after receiving vaccine, using an ELISA platform ^8^. These results demonstrated that a single drop of fingerstick blood-based testing method is sufficient for quantitative detecting and monitoring the development of anti-SARS-CoV-2 Spike IgG antibody after vaccination.

Notably, this assay is specific for vaccine elicited anti-SARS-CoV-2 S1 IgG antibody and does not reactive to anti-SARS-CoV-2 nucleocapsid (N) antibody, allowing us to distinguish vaccination from infection because SARS-CoV-2 S1 mRNA vaccine only stimulates anti-SARS-CoV-2 spike IgG antibody response but not anti-SARS-CoV-2 N protein IgG antibody. This will be useful with identifying vaccinated idividuals from infected idividuals and recovered patients if we apply two tests (two wells of 96 wellplate) with S1 coated beads and N-coated beads separately for each individual sample.

We tested more than 50 voluntary subjects in this study and observed no detectable anti-SARS-CoV-2 IgG before vaccination and during the first week among inviduals who had not been previously diagnosed, whereas the antibody became detectable 2 weeks after the first dose, averaging 5000 MFI. The antibody response increased to about 12000-13000 MFI by 7-8 weeks (i.e., 3-4 weeks after second vaccine dose, Figure 3). Notably, one individual infected with COVID-19 in 2020 whose IgG signal intensity jumped from 1300 MFI to over 15000 MFI whitin 1 week after first dose vaacination. This confirm seropositive persons has higher respond after first dose ^8^. Marot et al ^19^ reported that anti-S IgG and anti-RBD IgG antibody levels in previously confirmed COVID-19 patients did not change significantly between 3 week and 3 months, despite a slight increase at 2 months. Based on our data, the vaccine-induced IgG antibody responses appear to follow the same course. In addition, SARS-CoV-2 neutralization (NT50) assay has demonstrated significant correlation with all antibodies measured by immunoassays IgGs including anti-S-BRD IgG ^20^. Taken together, high levels of anti-SARS-CoV-2 spike IgG antibodies likely imply protective immunity against SARS-COV-2.

We found that all 3 subjects with undetectable anti-S IgG 2, two weeks after vaccination, the first vaccine dose were samples from over 80 years of age old. This observation suggests that not all vaccinated people will have detectable humoral immune responses within 2 weeks of the first vaccine dose, especially in the elderly population. However, all voluntary subjects including the elderly developed robust IgG antibody responses after the second dose of mRNA vaccine. Nevertheless, it should be noted that none of the voluntary subjects were considered immunocompromised.

In conclusion, we demonstrated that a single drop of fingerstick blood collected onto flocked swab can be used for quantitative detection and monitoring of anti-SARS-CoV-2 Spike IgG responses after receiving COVID-19 vaccination. This methodology is appealing because it makes possible for vaccinated individuals to collect samples without visiting healthcare facilities or clinical laboratories.

## Data Availability

all data referred to in the manuscript are available for public

## Supplementary

**S Figure 1.**
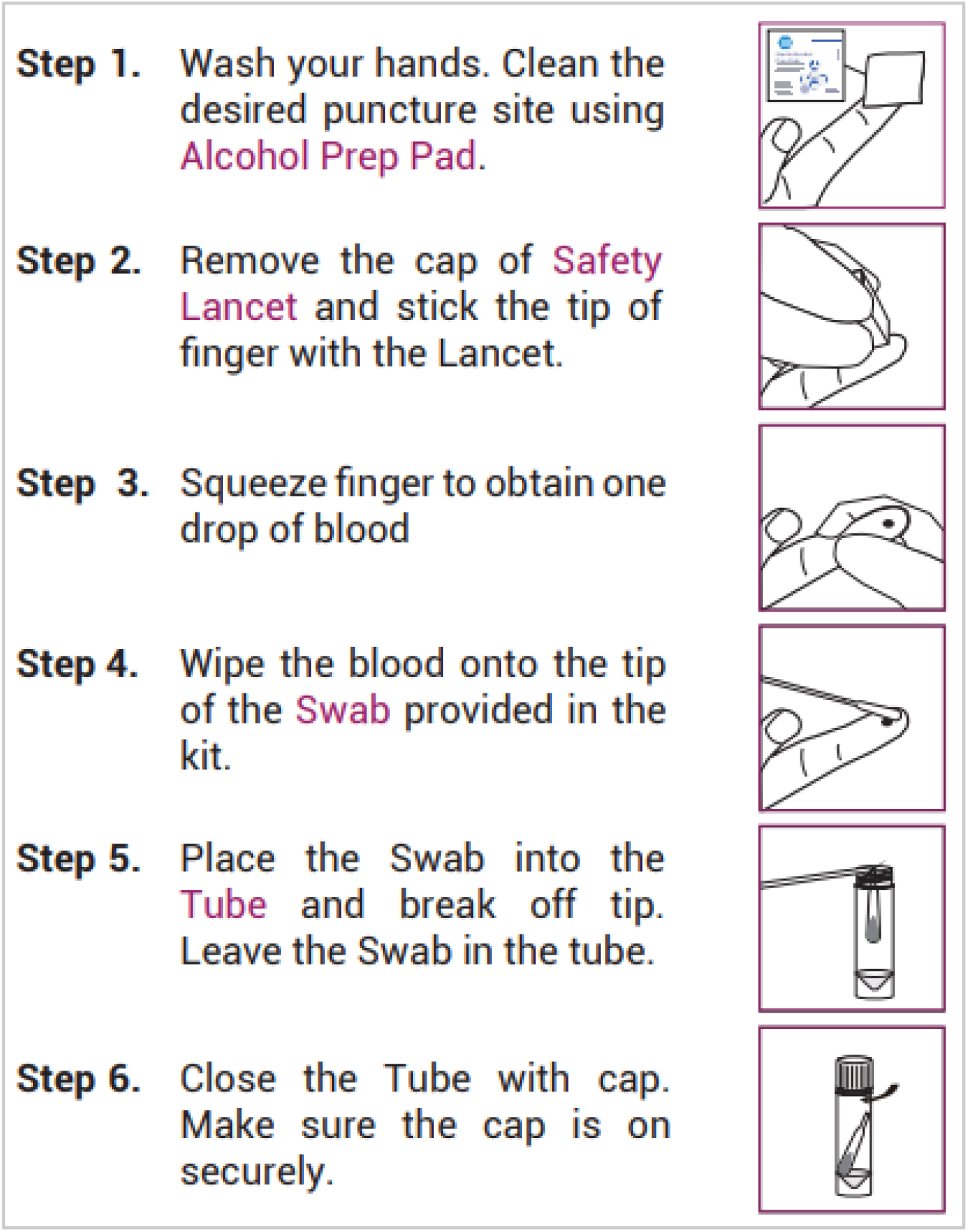
Fingerstick blood collection workflow

**S. Figure 2.**
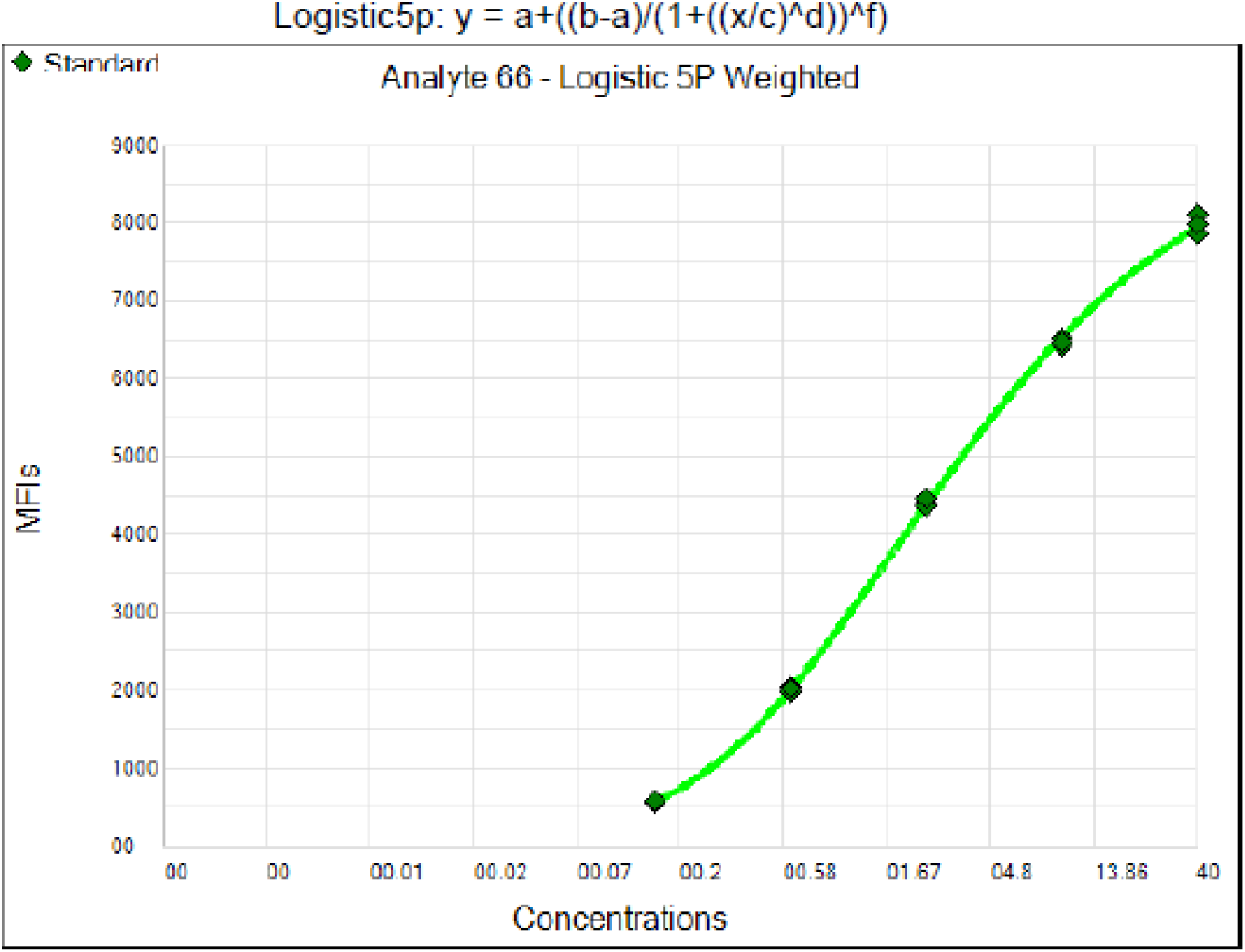
Standard curve generated by Luminex xPONENT software (R^2^ = 0.9996). X-axis: IgG concentration (μg/mL); Y-axis: Median Fluorescence Intensity (MFI).

